# Advance Transfer Learning Approach for Identification of Multiclass Skin Disease with LIME Explainable AI Technique

**DOI:** 10.1101/2024.12.02.24318311

**Authors:** Muhammad Zawad Mahmud, Md Shihab Reza, Shahran Rahman Alve, Samiha Islam, Nafis Fahmid

## Abstract

In dermatological diagnosis, accurately and appropriately classifying skin diseases is crucial for timely treatment, thereby improving patient outcomes. Our goal is to develop transfer learning models that can detect skin disease from images. We performed our study in the “Skin Cancer: MNIST HAM10000” dataset. This dataset has seven categories, including melanocytic nevi, melanoma, benign keratosis (solar lentigo/seborrheic keratosis), basal cell carcinoma, actinic keratoses, intraepithelial carcinoma (Bowen’s disease), vascular lesions, and more. To leverage pre-trained feature extraction, we use five available models—ResNet50, InceptionV3, VGG16, VGG19, and MobileNetV2. Overall results from these models show that ResNet50 is the least time-intensive and has the best accuracy (99%) in comparison to other classification performances. Interestingly, with a notable accuracy of 97.5%, MobileNetV2 also seems to be adequate in scenarios with less computational power than ResNet50. Finally, to interpret our black box model, we have used LIME as an explainable AI technique (XAI) to identify how the model is classifying the disease. The results emphasize the utility of transfer learning for optimizing diagnostic accuracy in skin disease classification, blending performance and resource efficiency as desired. The findings from this study may contribute to the development of automated tools for dermatological diagnosis and enable clinicians to reduce skin conditions in a timely manner.

## I. Introduction

Skin cancer represents the most common kind of cancer and includes types such as basal cell carcinoma, squamous cell carcinoma, and melanoma. In contrast to the considerably more malignant melanoma, it is far rarer, has a high metastatic potential, and is responsible for nearly all cancer deaths. It is one of the most common cancers in humans and among various human skin diseases. It is due to the effect of DNA injury, which can be induced by UV exposure from sunlight as well as other causes that create alterations to skin cell genes [1]. Risk factors for skin cancer include exposure to UV radiation, a light complexion, occupational exposures (farming), and genetic susceptibilities; these are believed to be some of the reasons why melanoma has become so common since the start of indoor tanning. The question of cancer is not only serious, but it is also responsible for a huge number of deaths linked to morbidity and mortality in association with this malignancy.

Skin cancer outranks all other malignancies in terms of morbidity, with a wide range of mortality rates both internationally and regionally. Bangladesh, for example, has acknowledged the impact more and is continually studying death rates to figure out what really is happening. On the flip side, until recently, skin cancer mortality rates in Bangladesh have been low compared to Western countries, but they are increasing as lifestyle and environmental exposures change is consistent with global research found elsewhere in the world linking skin cancer to an unacceptably high burden of mortality and morbidity, so similar investigations are being conducted as well beyond Australia (e.g., Asia) to capture this impact. However, skin cancer death rates reflect marked heterogeneity in both geographic and ethnic-racial origin here within the much broader Asian context. It also causes thousands of deaths annually worldwide (*>*1 million diagnosed cases per year).

The discovery of skin cancer is traditionally done by visual inspection and biopsy, where suspected lesions are analyzed to confirm malignancy. However, technological advancements, especially deep learning, have improved it to a new level altogether. This is made possible by the discriminative power of deep learning models that have been trained on large-scale datasets of skin images and provide high accuracy in detecting malignant features — a useful tool for early diagnosis as well as treatment planning. It recommends a clinical diagnosis, followed by dermoscopy and biopsy in detecting skin cancers. However, that has completely changed since deep learning appeared. For instance, deep learning models like convolutional neural networks (CNN) have achieved dermatologist-level performance in malignant skin lesion recognition. We decided to build the best results possible with regard to the classification of disease. The main contributions of our study are:

- Our main contribution is finetuning ResNet50 model to classify skin disease from the “Skin Cancer: MNIST HAM10000” dataset. This outperformed all existing accuracy models to this point and became the state-of-the-art model for this dataset.
- The second one is to apply LIME XAI to understand the ResNet50 model decision-making process.

## II. Related Work

The rapidly evolving field of computer-aided diagnostic technology has dramatically improved the ability to accurately diagnose skin cancer, which is essential due to the high death rates associated with the condition. Over the past few years, there have been amazing deep-learning methods that can help improve diagnostic results significantly for early detection, which leads to better treatment outcomes. This literature review aims at various types of research that used the “Skin Cancer: MNIST HAM10000” dataset to build various DL models for skin lesion classification. This review seeks to achieve a synthesis of these results, with emphasis on the operating characteristics regarding how effectively DL models distinguish malignant from benign skin lesions and what this means in clinical practice.

Sönmez et al. [2] classified that convolutional neural networks in deep learning are significantly important for classifying and detecting skin lesions accurately using dermoscopic images. This has been attributed to the fact that such authors primarily used the large-scale MNIST HAM10000 dataset with 10015 images for seven skin disorders. The authors further agree that the existing imbalanced data challenges present in such datasets can only be solved by using extensive data augmentation and profound, intelligent preprocess modifications. The paper illustrates an accuracy of 80.79% for the CNN classification task, whereas, in the MobileNetV2 model, the classification is disregarded.

Sandhua et al. [3], in their paper “Deep Learning-Based Skin Cancer Identification,” demonstrated the feasibility of CNNs to be applied in developing an approach for the independent categorization of skin cancers proactively, without human participation. In their research, they used the CNN model and trained them with the transfer learning models MobileNetV2 and ResNet 50, which provided excellent performance. Using that data, both of these models got the top accuracy with 96% and 89%.

Gururaj et al. [4] developed a deep learning technique in skin lesion diagnosis using CNNs, which achieves remarkable results. In comparison with existing data preprocessing, this work focuses on bettering the state-of-the-art models, such as DenseNet169 and ResNet50, which are known for outperforming architectures in the case of object detection. Apparently, these CNN models can give a significantly low error rate (91.2%) with the DenseNet169 model, and the F1 score is also remarkable—up to 91.7%) by just properly managing data using under-sampling and oversampling methods, as demonstrated in the research work.

Shamsi et al. [5] proposed a novel deep uncertainty diagnostic model based on Monte Carlo dropout (MCD) and entropy loss optimization to achieve assisted learning of skin cancer diagnosis. It is a significant development in the field since it overcomes some of the problems that have been found when implementing traditional MCD, particularly with respect to issues around or associated with overconfidence. The prediction accuracy of the built model is 85.65 ±0.18% on average.

The paper, titled “Skin lesion classification on dermatoscopic images using effective data augmentation and a pretrained deep learning approach” by Bozkurt [6], introduces a novel transformation that includes resampling methods that are done in different sizes using pre-trained deep-learning models. He proposes a unique technique to magnify the dataset using affine transformation techniques, which may help in improving completions. The accuracy results are pretty impressive: 95.09% accuracy with Inception-Resnet-v2.

Arun and Palmer [7] have assessed the suitability of applying CNN in automatic detection systems for skin cancer analysis from dermatoscopic images. They worked to develop a CNN model that analyzes transformed images in order to clearly highlight features typical of benign and malignant skin lesions. The model gave an accuracy of 95%.

Garg et al. [8] contracted a deep learning-based decision support system for the identification and classification of skin cancer using a combined CNN approach. The diagnosis is based on deep learning and image processing models. Their transfer learning model, ResNet50, reported the highest accuracy at 90.5%.

Alam et al. [9] unveiled a deep learning-based classifier for skin cancer as well. This paper proposes a groundbreaking automatic skin cancer detector based on deep learning using an unbalanced dataset. RegNetY-320 performed best among their applied models with 91% accuracy.

Cengil et al. [10] built hybrid CNN models for skin cancer classification. KNN, SVM, and DT are utilized in the realization of hybrid architectures. Following CNN feature extraction, KNN, SVM, and DT are used independently for classification. Alexnet+SVM gave the maximum accuracy (77.8%).

From the above discussion, it is clear that our model performed better than the existing studies of the dataset. Our novelty lies in introducing the LIME explainable AI method for this dataset, which no one integrated earlier.

## III. Methodology

### A. Dataset

The data for this paper was obtained from the Kaggle dataset. Data classes are classified into seven types. Those are melanocytic nevi, melanoma, benign keratosis, basal cell carcinoma, actinic keratoses, intraepithelial carcinoma, vascular lesions, and dermatofibroma. These are labeled as 0 to 6 for generating results. Melanocytic nevi have 6705 samples, melanoma has 1113 samples, benign keratosis has 1099 samples, basal cell carcinoma has 514 samples, actinic keratoses, and intraepithelial carcinoma have 327 samples, vascular lesions have 142 samples, and dermatofibroma has 115 samples. This information is used to diagnose skin diseases. The data used to support the research outcome are freely available at [11], [12].

### B. Data Preprocessing

It is clear from the above discussion that this dataset is imbalanced. Data augmentation was applied to balance the dataset. Additional images have been created from the original picture using standard data augmentation techniques, including rotation (20 degrees), width and height shifts (0.20), shear (0.20), zoom (0.20), horizontal flips, and nearest fill mode using *ImageDataGenerator* framework [13].

### C. Training Methodology

This subsection explains how the models were trained for this study.

#### 1) ResNet50

The ResNet50 model with pre-trained weights is known for using residual connections to solve the difficulties of deep learning. As part of preprocessing, the images were resized to 224×224 pixels as ResNet50 expects these dimensions. The model was trained for 30 epochs with a batch size of 64. The model was fine-tuned with the Adam optimizer. The early stopping was used to monitor validation loss automatically. Along with that, the *ModelCheckpoint* was used, which will be helpful in saving the best model based on validation accuracy. Furthermore, *ReduceLROnPlateau* was used to reduce the learning rate if the validation performance did not improve.

#### 2) InceptionV3

The InceptionV3 model uses inception modules for multi-scale feature extraction, and the initial weights of ImageNet are loaded. This InceptionV3-specific preprocessing function will resize the input image to 224×224 pixels. The model was trained with 30 epochs and a batch size of 64. The training was performed on the Adam optimizer with early stopping based on validation loss improvements controlling training duration. Next, *ModelCheckpoint* was used to save the model with the best validation accuracy, and similarly, an in-built callback method called *ReduceLROnPlateau* was set up, which would decrease the learning rate if our model performance on val did not improve.

#### 3) VGG16

A deep architecture with small convolutional filters— VGG16 model with pre-trained weights were loaded. Resizing images to 224×224 pixels as required by the VGG16 preprocessor. The training process lasted 30 epochs and had a batch size of 64. Training was performed using the Adam optimizer, and early stopping stopped when validation loss did not improve. To save the model that achieved the best validation accuracy, The *ModelCheckpoint* was used, and for dynamic learning rate adjustment based on validation performance (plateauing of training), a callback *ReduceLROnPlateau* was defined.

#### 4) VGG19

The VGG19 model (which denotes an extended version of 16-layer VGG with extra layers) was set up as pretrained weights. The preprocessing was resizing input images to 224×224 pixels for the VGG19 model. The above model has been trained for 30 epochs using a batch size of 64. The training was early-stopped based on the validation loss, and the Adam optimizer was implemented. The *ModelCheckpoint* was used to save the best validation accuracy model, and the learning rate of the optimizer was adjusted using *tensorflow*.*keras*.*callbacks*.*ReduceLROnPlateau* based on the same above scenario.

#### 5) MobileNetV2

Releasing a pre-trained variant of the efficiency-focused MobileNetV2 model architecture, which is tuned to mobile phone CPUs by replacing Conv 1×1 with depthwise convolutions. This is needed because MobileNet-v2 images are of size 224×224 pixels. Training runs for 30 epochs with batch size = 64. The Adam optimizer was used to optimize the model and set up early stopping to stop running more epochs if the validation loss didn’t improve. Furthermore, the best-achieved model was saved according to validation, and *ReduceLROnPlateau*, a Keras class that just automatically updates the learning rate on the go while observing a diminution of performance during training, was applied.

### D. LIME

LIME (Local Interpretable Model-Agnostic Explanations) is model-agnostic, and it can interpret any black-box classifier. LIME is among the simplest and most common XAI techniques. This model agnosticism refers to the fact that LIME is explainable for any supervised deep learning models in this world. The LIME takes in a prediction model and test sample as input. First, it does the sampling to get a surrogate dataset. By default, it normalizes the project feature vector and creates 5000 pseudo-samples. This process results in the target variable of these 5000 samples being achieved using that predictive model. The surrogate dataset also weights each row as a function of how closely the newly created samples resemble the original sample [14].

### E. Testing Methodology

This subsection explains how the models were tested for this study. The equations of the testing metrics are given below [15]:

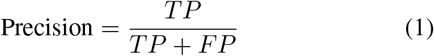

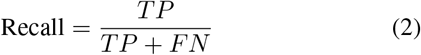

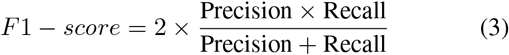

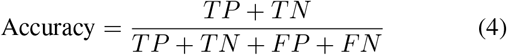

### F. Work Flow Diagram

Fig. 1 represents the workflow diagram of this study. Firstly, the dataset was loaded. Secondly, the data were preprocessed by augmenting and resizing. Thirdly, the dataset was split into an 80-20 ratio. 80% of the data was used to train the models (i.e., ResNet50, InceptionV3, VGG16, etc.). Finally, the models were evaluated on the remaining 20% of the data. Along with that, explainable AI (LIME) analysis was completed.

**Fig. 1.**
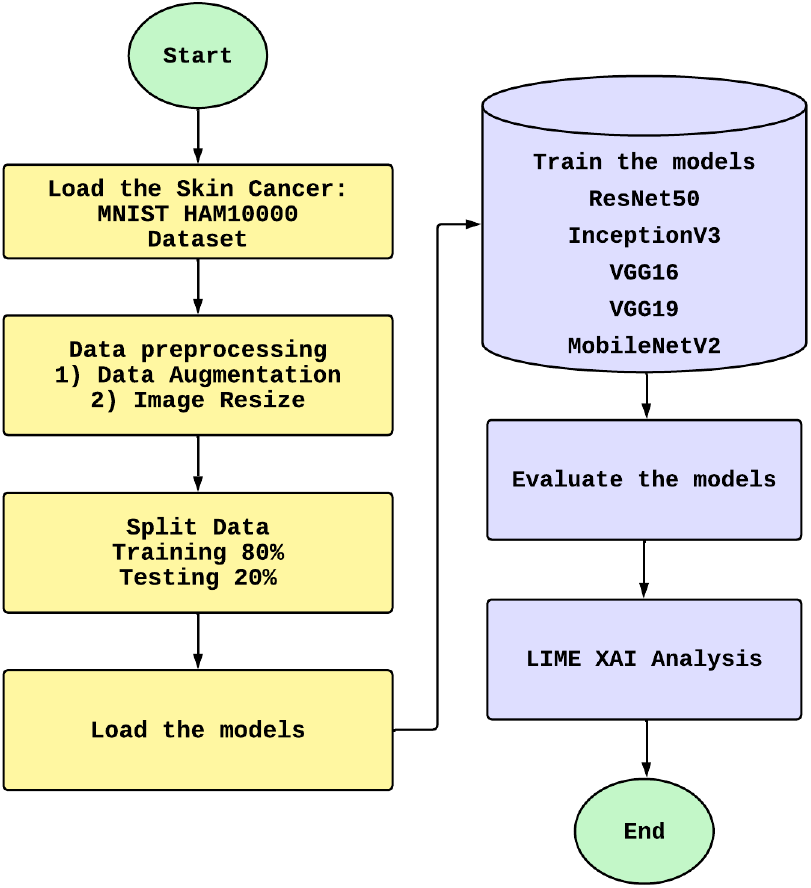
A detailed representation of the study’s workflow

## IV. Result Analysis

To classify skin cancer images in our study, we used five different CNN models. Optimized model wise: ResNet50, InceptionV3, VGG16, VGG19 and MobileNetV2. We evaluated these transfer learning models using five performance metrics, i.e., accuracy, precision, recall, f1-score, and confusion matrix. The lower subsection displays these metrics for the best and lower-performing models.

### A. ResNet50

Fig. 2 shows the accuracy curves of the ResNet50 model applied to the monochromatic image dataset of this study and classified as depicting non-reference radiographs. The trend-line of both accuracy on training and testing is continuous, while the excess of test over train is even better than expected during almost all epochs. By the 30th epoch, both accuracies converge close to 1.0, showing that your model now has high accuracy in the training and testing phases as well.

**Fig. 2.**
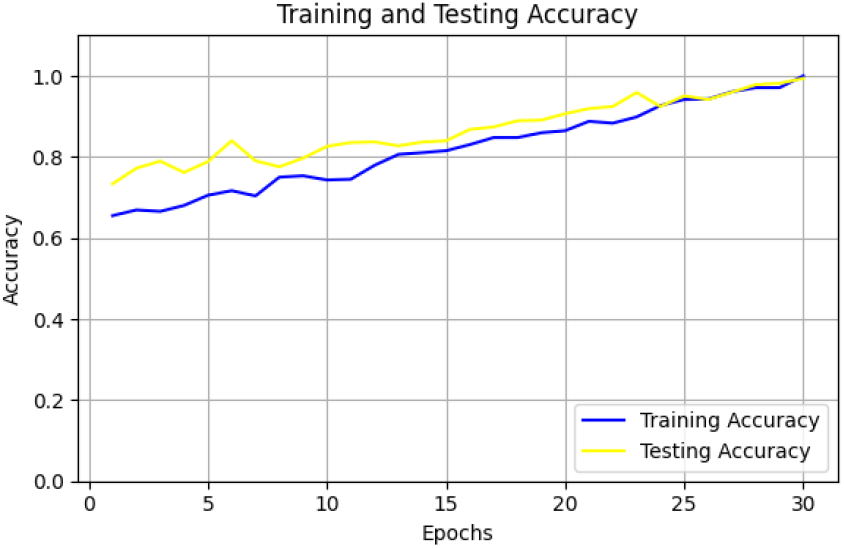
Train and test accuracy curve of ResNet50

The training and testing curves for the ResNet50 models of this study are depicted in Fig 3. The training loss and testing losses both decrease through iterations. The training loss is always less than a target, and when the run is over, all losses are equivalent to 0, which means that by one measure or another, the model works at full capacity.

**Fig. 3.**
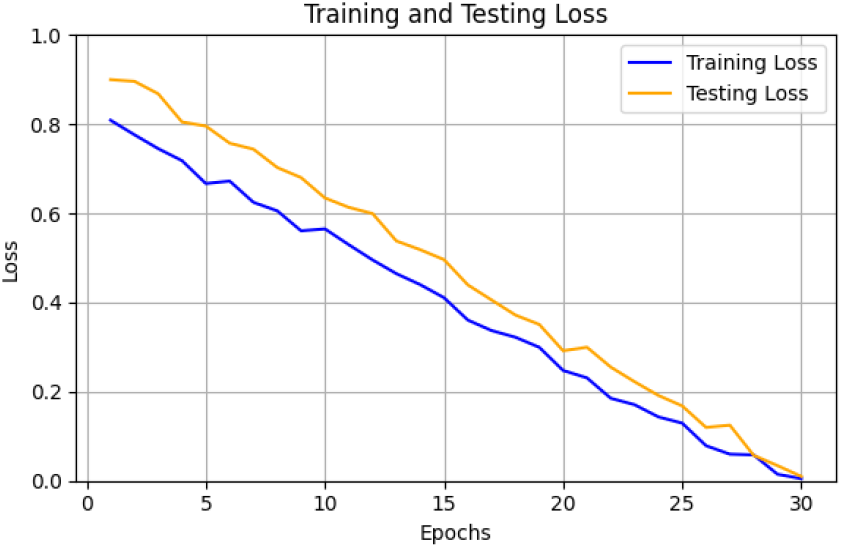
Train and test loss curve of ResNet50

Fig. 4 visualizes the confusion matrix for the ResNet50 model. ResNet50 exhibits superior performance across all classes, attaining near-perfect accuracies—99% for classes 0, 3, and 5, and a flawless 100% for class 1. Even in the previously challenging classes 2 and 6, ResNet50 records impressive accuracies of 98%, respectively.

**Fig. 4.**
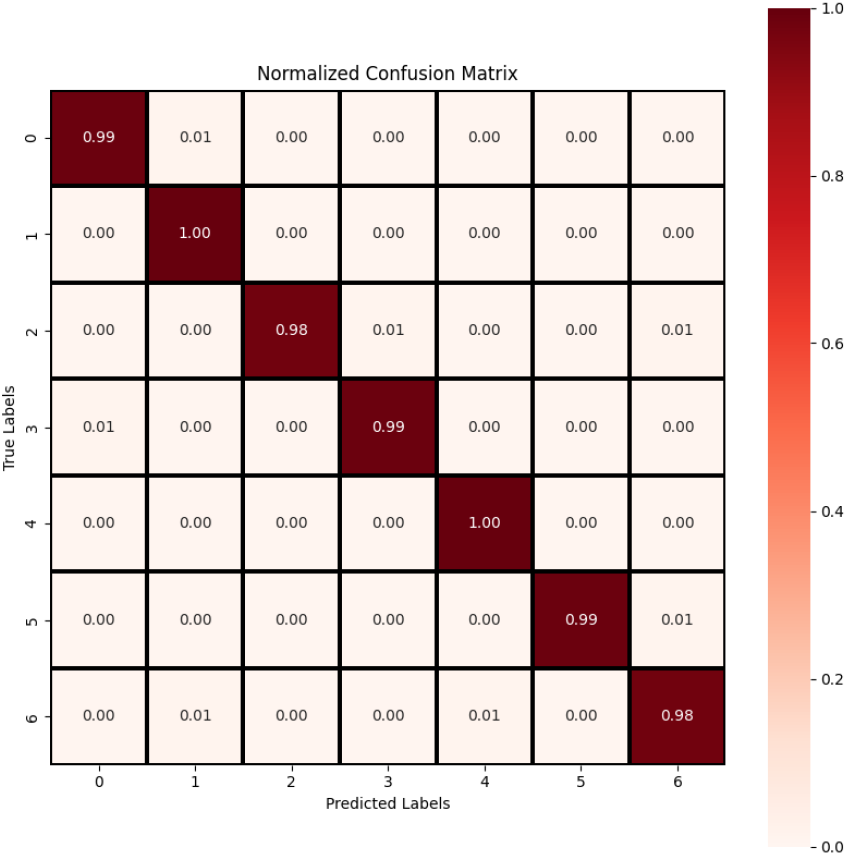
Confusion Matrix of ResNet50

### B. InceptionV3

Fig. 5 refers to the training and testing accuracy curve of the InceptionV3 model. The training accuracy (blue curve) constantly improved and reached 85% at the end of training. This trend is followed by the testing accuracy too (yellow curve), and it goes to 83.2% at the end of training. These values for accuracy are close, meaning that our model is not only suitable during the training but also during the testing phase.

**Fig. 5.**
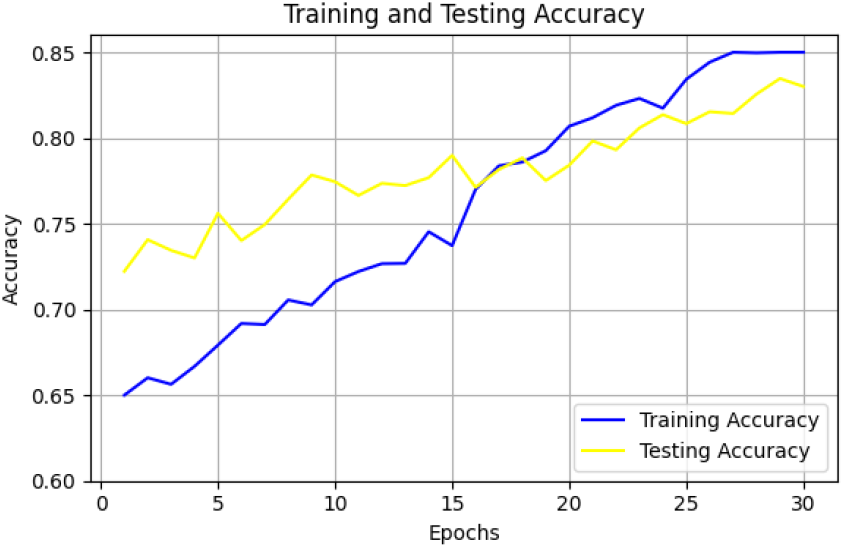
Train and test accuracy curve of InceptionV3

In Fig. 6, the InceptionV3 model’s training and testing loss curves are displayed. The plots show how both the losses decrease steadily over 30 epochs. Even as training progresses, the training loss is consistently less than the test loss. The two losses converge at 0.10 during the last epoch, where they remain until training finishes, meaning that our model has effectively minimized error during training phases as well.

**Fig. 6.**
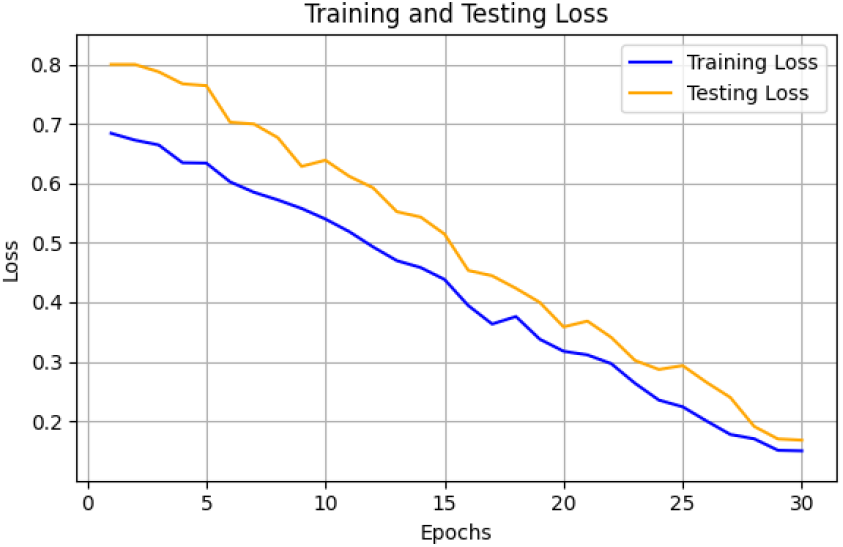
Train and test loss curve of InceptionV3

Fig. 7 shows the confusion matrix of InceptionV3. The model achieves 91% accuracy for categories (0, 1), but it loses accuracy over categories, reaching 88% for category 2, then drops to around only 83% and down to just above the chance (81%) of correct classification across our broadest genre definition. It is difficult because, with instances of categories 5 and 6, its accuracies drop to as low as 75% or even lower, for example, at 68%.

**Fig. 7.**
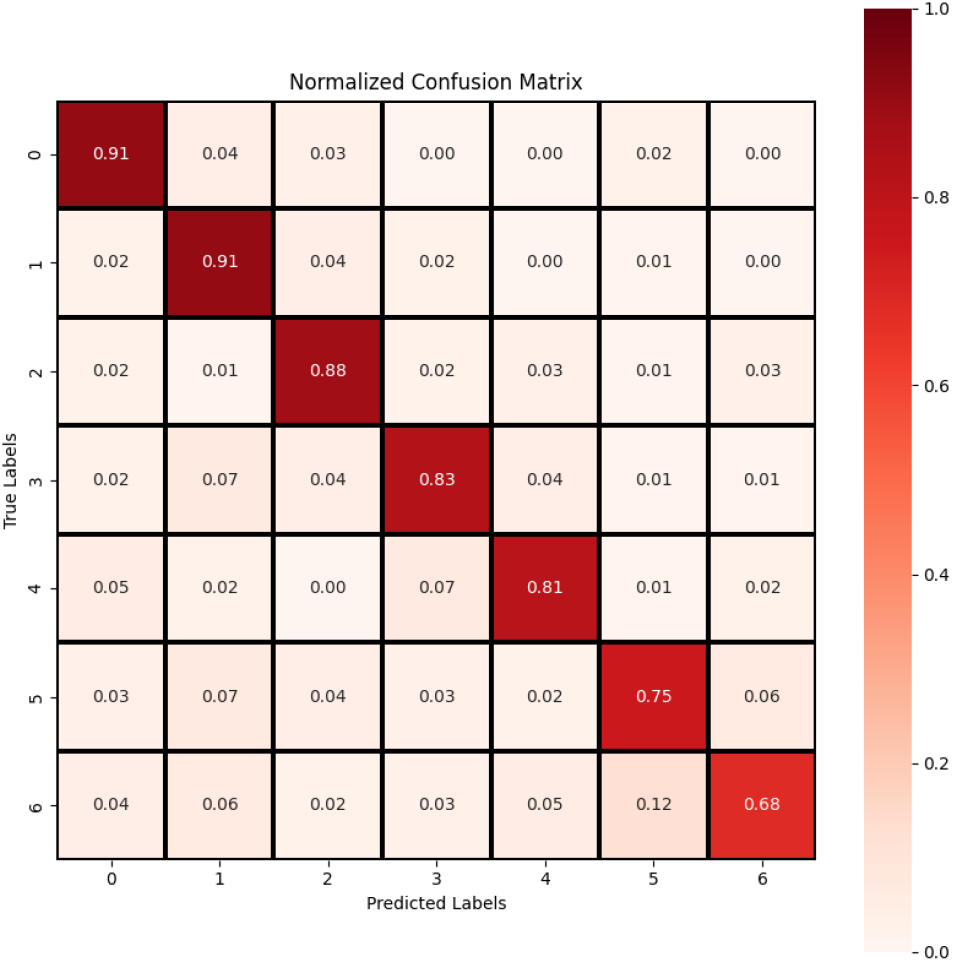
Confusion Matrix of InceptionV3

### C. Model Evaluation

Performance results from the transfer learning models are presented in Table I, where ResNet50 shows good performance over all the models with a test accuracy of 99% and a low value for test loss of 0.08, which highlights its potential generalization capabilities as well.

**TABLE I.** Result Evaluation of Models.

| Models | Train accuracy (%) | Test accuracy (%) | Test Loss |
| --- | --- | --- | --- |
| ResNet50 | 99.58 | 99.00 | 0.010 |
| InceptionV3 | 85.00 | 83.00 | 0.168 |
| VGG16 | 88.20 | 86.00 | 0.130 |
| VGG19 | 89.50 | 87.30 | 0.132 |
| MobileNetV2 | 98.20 | 97.50 | 0.018 |

Table II presents parameters in millions, precision, recall accuracy, and F1-score of the used models. We got the best results with ResNet50 in all metrics, which also had a precision and recall of 0.99 and an AUC equal to 0.99. It shows how strong ResNet50 is when it comes to classifying, which results in the clean classification of types or groups of skin cancer with the least FP and FN, coming on top performance.

**TABLE II.** Performance Metrics Comparison.

| Models | Parameters | Precision | Recall | F1-Score | AUC |
| --- | --- | --- | --- | --- | --- |
| ResNet50 | 25.6M | 0.99 | 0.99 | 0.99 | 0.99 |
| InceptionV3 | 23.9M | 0.81 | 0.79 | 0.80 | 0.85 |
| VGG16 | 138.4M | 0.87 | 0.85 | 0.86 | 0.88 |
| VGG19 | 143.7M | 0.87 | 0.85 | 0.86 | 0.88 |
| MobileNetV2 | 3.5M | 0.96 | 0.95 | 0.96 | 0.98 |

### D. Result Comparison

As shown in Table III, the models are compared to those previously studied in the “Skin Cancer: MNIST HAM10000” dataset. According to the table, the ResNet50 model outperforms all others in the framework. MobileNetV2 also demonstrated superior performance, albeit with significantly fewer computational resources than ResNet50.

**TABLE III.** Result Comparison.

| Study | Model | Accuracy (%) |
| --- | --- | --- |
| This study | ResNet50 | **99.00** |
| This study | MobileNetV2 | **97.50** |
| Sandhua et al. [3] | MobileNetV2 | 96.00 |
| Bozkurt [6] | Inception-Resnet-v2 | 95.09 |
| Arun and Palmer [7] | CNN | 95.00 |
| Alam et al. [9] | RegNetY-320 | 91.00 |

### E. Explainable AI

From our study, ResNet50 emerged as the best-performing model, so we applied the LIME XAI with the ResNet50 model. We have checked random images from our dataset to explain and show that the regions that determine the predictions made by ResNet50 can be visualized., which is shown in Fig. 8. The leftmost images of the figures depict a benign keratosis-like lesion (BKL) and melanoma (MEL). The middle row gives the segmentation mask representing different regions from which the model learns per each provided image. The dark region in the mask represents more critical regions, showing that it leads to a low value. Areas with higher intensity in the mask indicate regions with greater influence on the model’s prediction. As shown in the figure, image of the rightmost supply attention to areas and contents fortunate enough to be attracted by our model. This indicates that texture and color in these regions are important for distinguishing between skin disease images. By highlighting sections in the image, LIME XAI helps us grasp cues and patterns that impact how the ResNet50 deep learning model categorizes inputs.

**Fig. 8.**
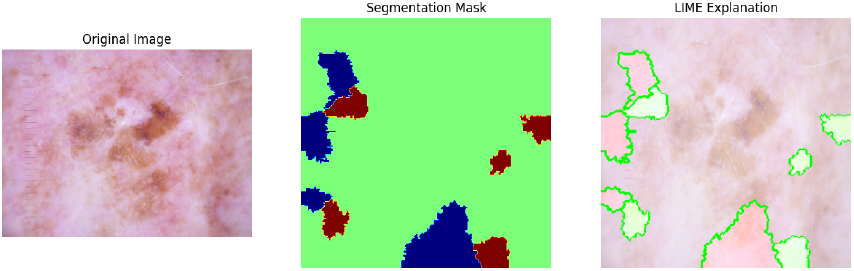
LIME XAI visualization for the ResNet50 model.

## V. Conclusion and Future Work

As an outcome, this paper exhibits the capability of transfer learning models on a multi-class skin disease classification task compatible with the dataset “Skin Cancer: MNIST HAM10000”. The results of the five models showed that ResNet50 had 99% specificity, which was the highest among all other models, and hence, it became the most useful model for this purpose. Furthermore, MobileNetV2 also showed good performance with an accuracy of 97.5%, demanding less computational resources as compared to the other models. The Diebold performance of these models demonstrates that transfer learning may be a promising way to become aware of skin diseases inside clinical pictures. In the future, we plan to further improve skin cancer diagnosis by introducing a novel implicit attention mechanism integrated into our hybrid CNN for small datasets. Exploring other XAI techniques like Grad-CAM for different models. One primary area where real-world data will be collated from Bangladeshi hospitals and the hybrid dataset created using this image data, which shall make model predictions more accurate and contextually relevant.

## Data Availability

All data produced in the present study are available upon reasonable request to the authors.

